# Leveraging HCUP Data to Predict Preventable Admissions in Patients with Chronic Kidney Diseases

**DOI:** 10.64898/2026.11.23.26368662

**Authors:** Yu-Chun Huang, Yuanji Han, Yun-Chung Liu, Chirag Parikh, Hadi Kharrazi, Derek Baughman

## Abstract

**Objectives:** This study developed machine learning (ML) to predict 90-day preventable admissions in CKD patients while evaluating racial disparities in model performance.

**Materials and Methods:** Study population included CKD patients (≥18 years) with hospital encounters in Maryland (2016–2023). Logistic regression (LR), random forest (RF), XGBoost, and k-nearest neighbors (KNN) models were used to predict 90-day preventable admissions after a CKD outpatient visit. Data used in this study was extracted from Healthcare Cost and Utilization Project (HCUP).

**Results:** 28,214 unique patients were included, of whom 3974 (14%) experienced a subsequent preventable admission. LR achieved comparable performance with RF and XGBoost (AuROC 0.784 - 0.788, AuPRC 0.397 - 0.416), outperforming KNN (AuROC 0.753, AuPRC 0.342). All models performed marginally better in the Black population (AuROC 0.773 - 0.807, AuPRC 0.369 - 0.467) than in the white population (AuROC 0.738 - 0.776, AuPRC 0.329 - 0.394). The false negative rates (FNR) in the white population were higher across different models (Black: 0.236 - 0.33, white: 0.297 - 0.377).

**Discussion:** Our results suggest that LR can achieve comparable prediction performances than tree-based ML algorithms. Hospital utilization and congestive heart failure are among the most important predictors across models. Further analyses on prediction bias revealed that FNRs are more significant in white patients, which can be due to their overall lower utilization.

**Conclusions:** This study demonstrated a pipeline to develop ML models predicting preventable admissions while revealing existing racial biases. These findings can inform ambulatory settings on how to reduce preventable admissions among CKD patients.

## Background and significance

Chronic kidney disease (CKD) impacts more than 10 % of Americans[1–3]. CKD complications include cardiovascular diseases and end-stage renal disease (ESRD). These conditions can impose significant economic burden and high hospitalization rates especially in disadvantaged populations. Hospitalization rate of patients with CKD was reported two-fold higher than the general population[2]. CKD hospitalizations due to ambulatory care-sensitive conditions, such as hyperkalemia, malignant hypertension, and heart failure[4], if managed adequately, could be preventable[5]. Prior population-based research has shown that CKD-related potentially preventable hospitalizations are concentrated among a relatively small subset of high-utilizing patients, suggesting the importance of identifying patients at elevated risk before preventable acute care use occurs[6].

Machine learning (ML) algorithms have shown considerable success when applied to CKD diagnosis, prognosis prediction, and treatment support[6]. For example, numerous ML algorithms have been used for CKD detection (e.g., support vector machine, deep neural network)[7], predicting CKD progression to ESRD (e.g., long-short term memory)[8], and predicting length of stay and associated costs due to CKD (e.g., random forest)[9]. ML algorithms have also been applied to predict hospitalizations due to CKD: predicting 90-day risk of hospitalization using claims data among Medicare patients[10], as well as predicting annual preventable hospitalizations in the Danish population[11]. However, none of these studies have attempted to predict preventable hospitalizations using sub-annual timeframes.

Despite the promise of ML in facilitating clinical decision making, research has shown that ML models can exacerbate, instead of mitigating, existing disparities in health utilization prediction[12–14]. Nonetheless, few ML studies focusing on CKD outcomes address potential biases across subpopulations such as racial groups or social economic strata and the results remain mixed[6, 15]. A study conducted on patients at Mass General Brigham showed that CKD patients differed in social determinants of health, such as insurance type and education level, in different racial groups. However, ML models showed no significant differences on mortality prediction across racial groups[16]. In another study, ML models were trained to predict five-year kidney-failure in CKD patients. The results suggest that even though the predictive accuracy was high, the error rate differed significantly across racial groups[17].

### Objective

To address the gaps in using ML for CKD outcome predictions, this study aims to develop ML models using population-level hospital discharge data to predict 90-day preventable emergency department (ED) and inpatient admissions due to CKD. Additionally, potential bias of these ML models across racial groups will also be assessed. The findings can be informative for preventive care measures in ambulatory care settings.

## Methods

### Data Source

This study used encounter-level data from the Healthcare Cost and Utilization Project (HCUP) database in Maryland from 2016 to 2023. The HCUP databases contain hospital discharge patient information from non-federal hospitals in the United States[18]. HCUP databases used in this study included datasets covering ambulatory visits, emergency department (ED) visits, and hospital admissions. HCUP data are deidentified and exempt from Institutional Review Board (IRB) reviews.

### Study Population

The study population of 28,214 patients included adult patients (≥ 18 years) diagnosed with CKD. CKD visits were defined as ambulatory encounters (Maryland, 2016-2023) with ICD-10 N18* diagnosis codes. Additional inclusion criteria included established patient visits (CPT 99211-99215) and one of the following confirmatory lab exam records (defined using CPT codes): creatinine (82565), basic metabolic panel (BMP, 80048), or comprehensive metabolic panel (CMP, 80053) within one year prior to the CKD visit[19], which improve the reliability of the CKD cohort and analysis[20].The exclusion criteria were end-stage renal disease (ESRD, N18.6), dialysis (Z99.2), kidney transplants (Z94.0), and palliative care (Z51.5)[21, 22].

### Outcome

The outcome of interest was preventable CKD admissions, defined as ED visits or hospitalizations attributable to heart failure (I50*), hypertensive crisis (I16*), hyperkalemia (E87.5*), or volume overload (E87.7*, R60.1, J81*) occurring within a 90-day period of the CKD visit[4, 20].

### Predictors

Predictors included demographic variables (i.e., age, sex, race, primary insurance payer), utilization (i.e., number of ambulatory visits, ED visits, or inpatient admissions within one-year prior to the CKD visit), and comorbidities (e.g., liver disease, diabetes, hypertension)[23, 24]. Predictors also included the disease groups derived from the Charlson Comorbidity Index (CCI), an assessment tool designed specifically to predict long-term mortality[25, 26], non-overlapping with comorbidities mentioned above (e.g., myocardial infarction, peripheral vascular disease) defined using ICD codes. Additional details of the predictors can be found in **Supplementary Table 1.**

### Model Selection

Logistic Regression (LR) and three other supervised ML models were developed to predict 90-day preventable CKD admission: Random Forest (RF), XGBoost, and K-Nearest Neighbors (KNN). LR was used as the baseline model due to its widespread use in clinical risk prediction and its interpretability, allowing estimation of the association between predictors and readmission risk[27]. RF and XGBoost were used to capture potential nonlinear relationships and complex feature interactions, which are known for their robustness and strong performance in structured healthcare data[9]. KNN was included as a distance-based, non-parametric method to provide contrast with both linear and tree-based approaches[7].

### Hyperparameter Optimization

Grid search and Bayesian optimization (using Optuna 4.9.0 Python package) with 30 trials per model were adopted to identify the best combination of hyperparameters[28, 29]. Both optimization procedures were repeated ten times using 3-fold cross validation. For each model, the hyperparameter combination achieving the highest AuROC across repetitions was selected. The complete set of selected hyperparameters for all four models under both grid search and Bayesian optimization is reported in **Supplementary Table 5.**

### Model Interpretability

SHAP(SHapley Additive exPlanations) analysis to LR, RF, and XGBoost using the Python *shap* library (version 0.51.0) for interpretability analysis. A representative model for each algorithm was selected based on the bootstrap iteration with AuROC was closest to the median across 100 splits. For tree-based models, TreeExplainer with path-dependent approximation was used to reduce the computational cost while maintaining result consistency with exact SHAP values for feature ranking process. For LR, LinearExplainer was applied, and the SHAP values were derived from the model coefficients directly.

To ensure stable coefficient estimation in LR and reliable feature contribution in SHAP across LR, RF, and XGBoost. Multicollinearity among predictors was assessed using the Variance Inflation Factor (VIF) prior to model training. Features with VIF exceeding 10 were removed. The VIF result is reported in **Supplementary Table 6.**

### Statistical Analysis and Model Evaluation

Characteristics of the patients experiencing the outcome of interest (90-day preventable CKD admissions) versus patients who did not experience the outcome were compared using chi-squared and t-test using the python *tableone* library (version 0.9.6). The dataset was randomly split into training (80%) and validation (20%) datasets.

Area under the receiver operating characteristics curve (AuROC) and area under the precision-recall curve (AuPRC) were used to evaluate the overall model performances. Accuracy, precision, recall, F1 score, and F2 score were calculated using the threshold maximizing the Youden Index[26]. Confidence intervals were calculated using bootstrapping techniques on 100 random training-validation dataset splits. All ML models were trained on the training dataset and the performance metrics were calculated using the held-out validation dataset. Data preprocessing was conducted using the *pandas* (version 2.2.3) and *polars* (version 1.35.2) libraries, model training and evaluation were accomplished by *scikit-learn* (version 1.5.1) and *XGBoost* (version 3.1.3) libraries of the python programming language.

## Results

### Study Population

As shown in **Figure 1**, 3,745,583 unique patients were identified with ambulatory visits to non-federal hospitals in Maryland between 2016 and 2023. Among them, 42,988 patients met the inclusion criteria of CKD diagnosis with records of established patients and relevant laboratory records (i.e., creatinine, BMP, and CMP) within one year prior to the index ambulatory visit. After excluding patients with ESRD (n = 13,251), dialysis (n = 302), kidney transplantation (n = 847), and receipt of palliative care (n = 374), the final study population included 28,214 patients.

**Figure 1.**
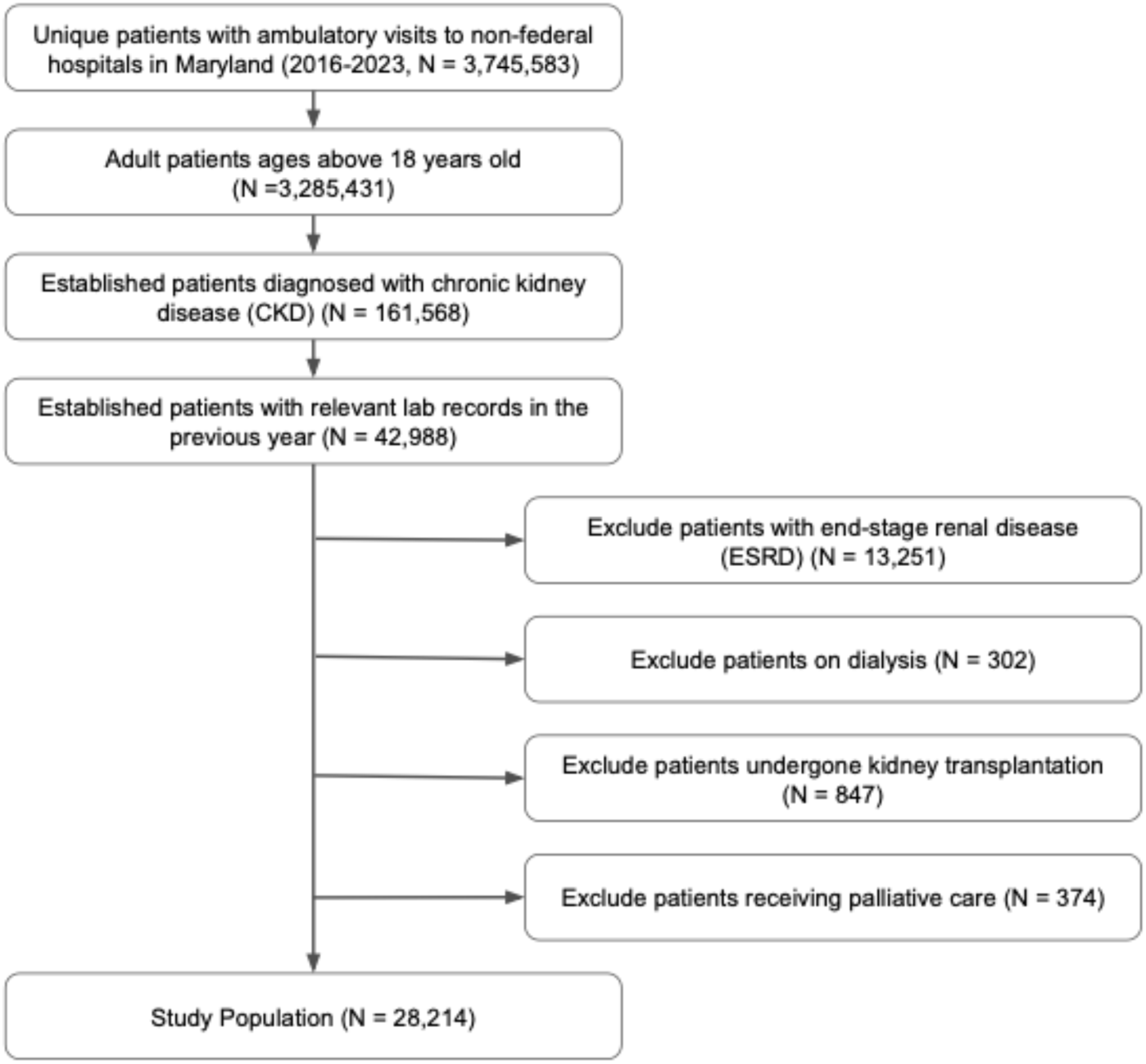
Flowchart of the study population.

**Figure 2.**
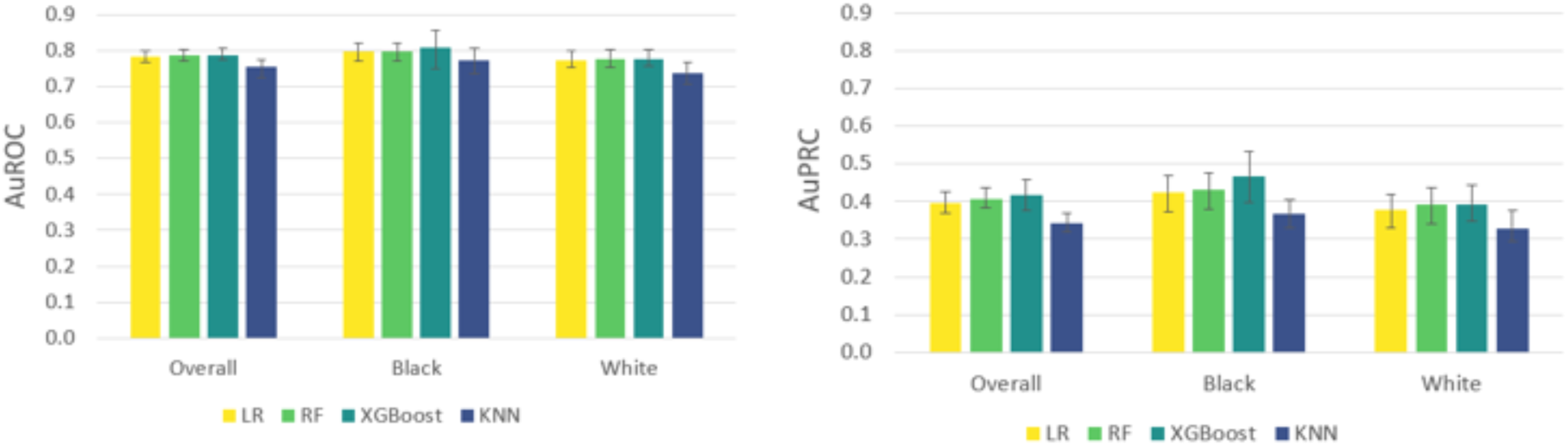
Model performance for predicting 90-day preventable CKD admissions using AuROC (left) and AuPRC (right)* * Bars represent mean performance across 100 random seeds, with error bars indicating 95% confidence intervals. LR: logistic regression, RF: random forest, KNN: k-nearest neighbors

**Figure 3.**
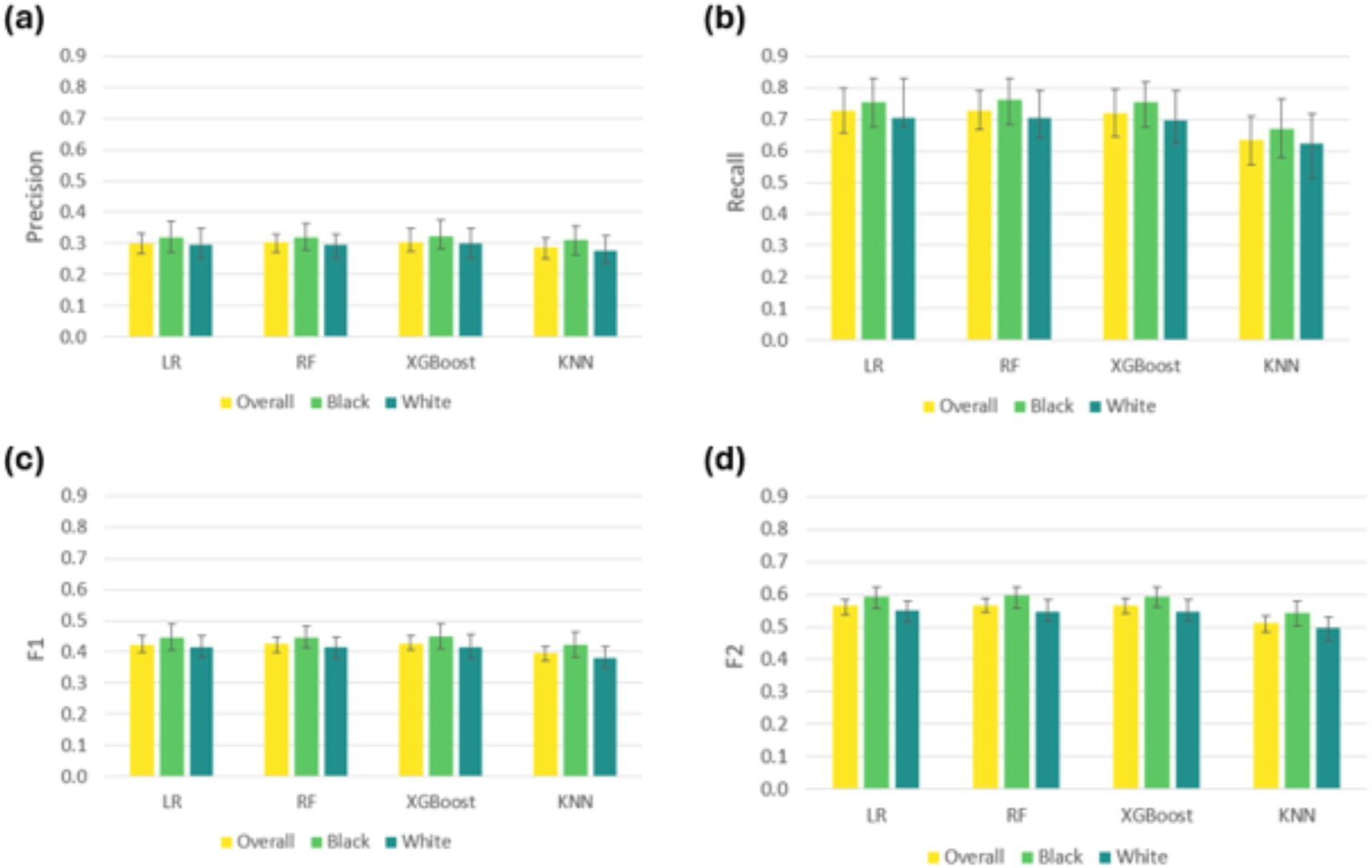
Model Performance Maximizing Youden Index measured by precision (a), recall (b), F1 score (c), and F2 score (d) *. *Bars represent mean performance across 100 random seeds, with error bars indicating 95% confidence intervals. LR: logistic regression, RF: random forest, KNN: k-nearest neighbors

### Patient Characteristics

As shown in **Table 1**, among the 28,214 included patients, 3974 experienced the outcome of interest of preventable 90-day CKD admission. Patients with the outcome were older on average than those without the outcome (70.8 ± 12.5 vs 68.4 ± 13.5 years; P < .001). Racial composition also differed between patients with and without the outcome (P = .005), with a higher proportion of black patients in the outcome group (43.2% vs 40.8%) and a slightly lower proportion of white patients (51.7% vs 52.9%). Insurance type distributions also differed significantly between groups (P < .001), with Medicare patients representing a larger proportion of patients in the outcome group (73.9% vs 66.9%). In addition, patients with the outcome were found to have higher hospital utilization (P < .001).

**Table 1.**
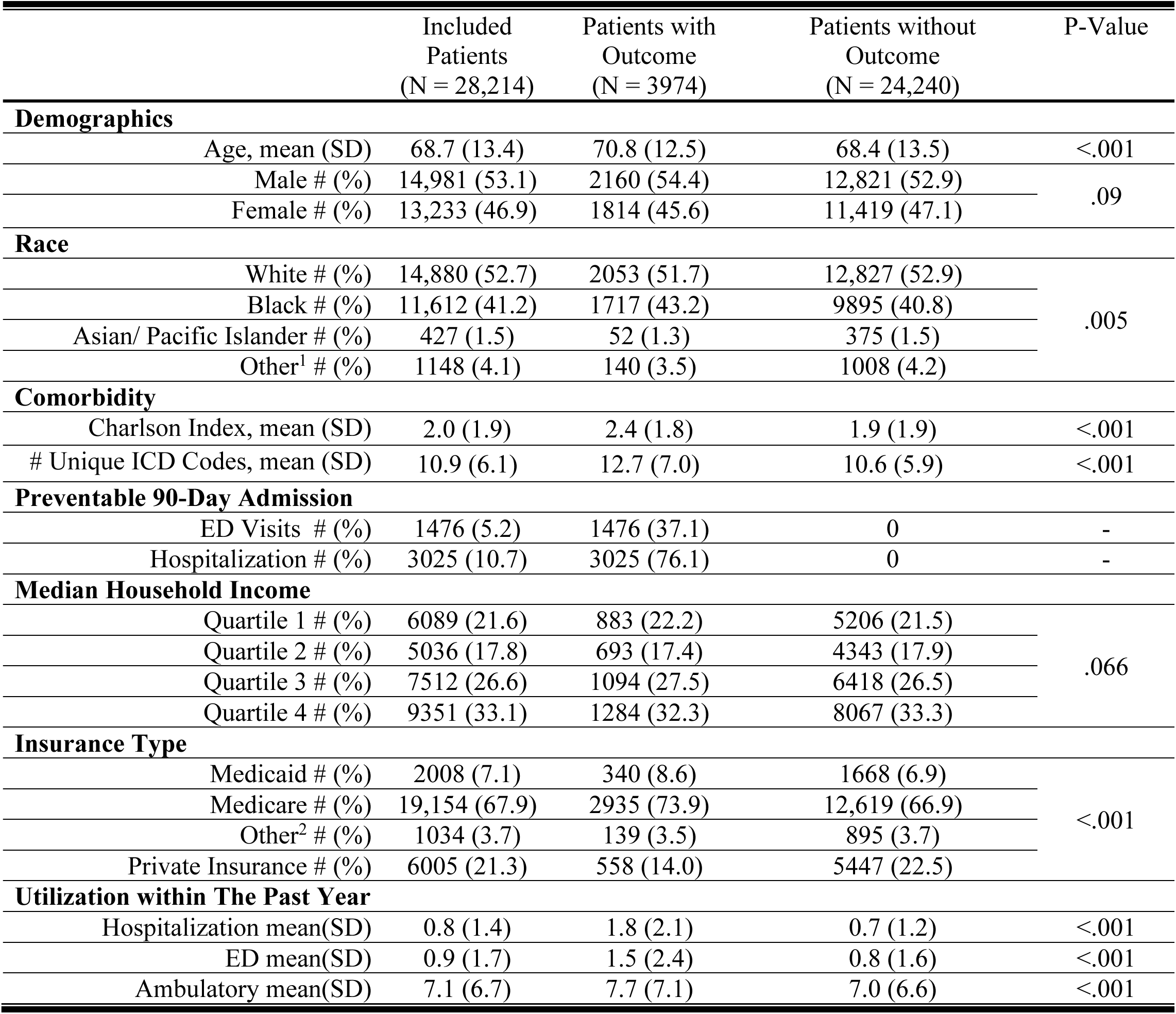
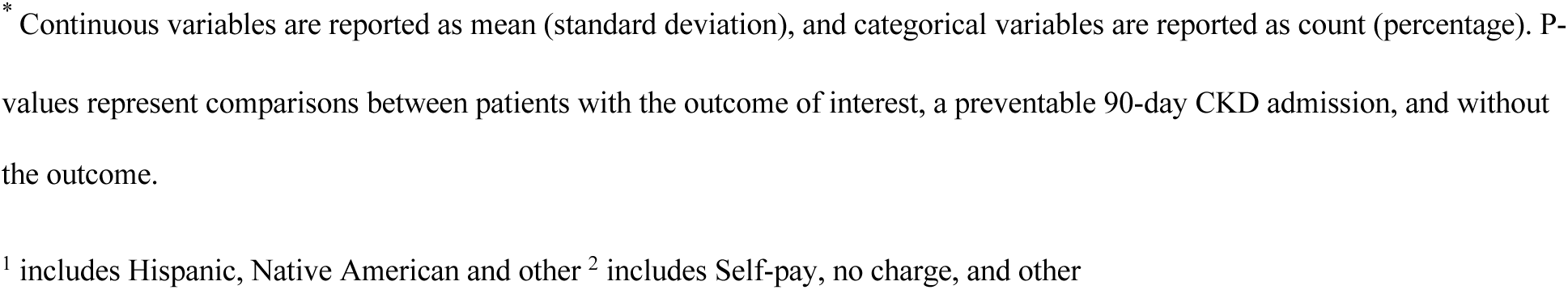
Characteristics of the Study Population*

|  | Included Patients<br>(N = 28,214) | Patients with Outcome<br>(N = 3974) | Patients without Outcome<br>(N = 24,240) | P-Value |
|---|---|---|---|---|
| **Demographics** |  |  |  |  |
| Age, mean (SD) | 68.7 (13.4) | 70.8 (12.5) | 68.4 (13.5) | <.001 |
| Male # (%) | 14,981 (53.1) | 2160 (54.4) | 12,821 (52.9) | .09 |
| Female # (%) | 13,233 (46.9) | 1814 (45.6) | 11,419 (47.1) |  |
| **Race** |  |  |  |  |
| White # (%) | 14,880 (52.7) | 2053 (51.7) | 12,827 (52.9) | .005 |
| Black # (%) | 11,612 (41.2) | 1717 (43.2) | 9895 (40.8) |  |
| Asian/ Pacific Islander # (%) | 427 (1.5) | 52 (1.3) | 375 (1.5) |  |
| Other<sup>1</sup> # (%) | 1148 (4.1) | 140 (3.5) | 1008 (4.2) |  |
| **Comorbidity** |  |  |  |  |
| Charlson Index, mean (SD) | 2.0 (1.9) | 2.4 (1.8) | 1.9 (1.9) | <.001 |
| # Unique ICD Codes, mean (SD) | 10.9 (6.1) | 12.7 (7.0) | 10.6 (5.9) | <.001 |
| **Preventable 90-Day Admission** |  |  |  |  |
| ED Visits # (%) | 1476 (5.2) | 1476 (37.1) | 0 | - |
| Hospitalization # (%) | 3025 (10.7) | 3025 (76.1) | 0 | - |
| **Median Household Income** |  |  |  |  |
| Quartile 1 # (%) | 6089 (21.6) | 883 (22.2) | 5206 (21.5) | .066 |
| Quartile 2 # (%) | 5036 (17.8) | 693 (17.4) | 4343 (17.9) |  |
| Quartile 3 # (%) | 7512 (26.6) | 1094 (27.5) | 6418 (26.5) |  |
| Quartile 4 # (%) | 9351 (33.1) | 1284 (32.3) | 8067 (33.3) |  |
| **Insurance Type** |  |  |  |  |
| Medicaid # (%) | 2008 (7.1) | 340 (8.6) | 1668 (6.9) | <.001 |
| Medicare # (%) | 19,154 (67.9) | 2935 (73.9) | 12,619 (66.9) |  |
| Other<sup>2</sup> # (%) | 1034 (3.7) | 139 (3.5) | 895 (3.7) |  |
| Private Insurance # (%) | 6005 (21.3) | 558 (14.0) | 5447 (22.5) |  |
| **Utilization within The Past Year** |  |  |  |  |
| Hospitalization mean(SD) | 0.8 (1.4) | 1.8 (2.1) | 0.7 (1.2) | <.001 |
| ED mean(SD) | 0.9 (1.7) | 1.5 (2.4) | 0.8 (1.6) | <.001 |
| Ambulatory mean(SD) | 7.1 (6.7) | 7.7 (7.1) | 7.0 (6.6) | <.001 |

<sup>\*</sup> Continuous variables are reported as mean (standard deviation), and categorical variables are reported as count (percentage). P-values represent comparisons between patients with the outcome of interest, a preventable 90-day CKD admission, and without the outcome.
<sup>1</sup> includes Hispanic, Native American and other <sup>2</sup> includes Self-pay, no charge, and other

### Model Performance

The overall model performances measured by AuROC and AuPRC are shown in **Figure 2**. LR, RF, and XGBoost reached comparable performance measured by AuROC overall (0.784 - 0.788) and in different racial groups (Black: 0.773 - 0.807, white: 0.738 - 0.776). The same trend can be observed using the AuPRC as the evaluation matrix for model performance. Notably, all models performed marginally better in the Black population than in the white population. For example, the AuPRC of LR is 0.425 in the Black population and 0.378 in the white population. The detailed performance metrics can be found in **Supplementary Table 2**. Prediction performance on different time-frames can be found in **Supplementary Figure 1**.

**Figure 3** summarizes model performances at the threshold maximizing Youden index. Similar to the trend observed in **Figure 2**, LR, RF, XGBoost showed comparable performances in terms of F-1 Score, outperforming KNN (0.423-0.427 vs 0.394). All models showed marginally higher performances in the Black population than in the white population (F1 score 0.423-0.447 vs 0.382-0.416, F2 score 0.542-0.595 vs 0.496-0.549). The detailed performance metrics can be found in **Supplementary Table 3**.

### Bias Measures

False positive rate (FPR) and false negative rate (FNR) of models were measured across racial groups (**Figure 4**). All four models show slightly higher FNRs in the white population then in the black population (white: 0.297-0.377, Black 0.236-0.33). Detail bias metrics can be found in **Supplementary Table 4**.

**Figure 4.**
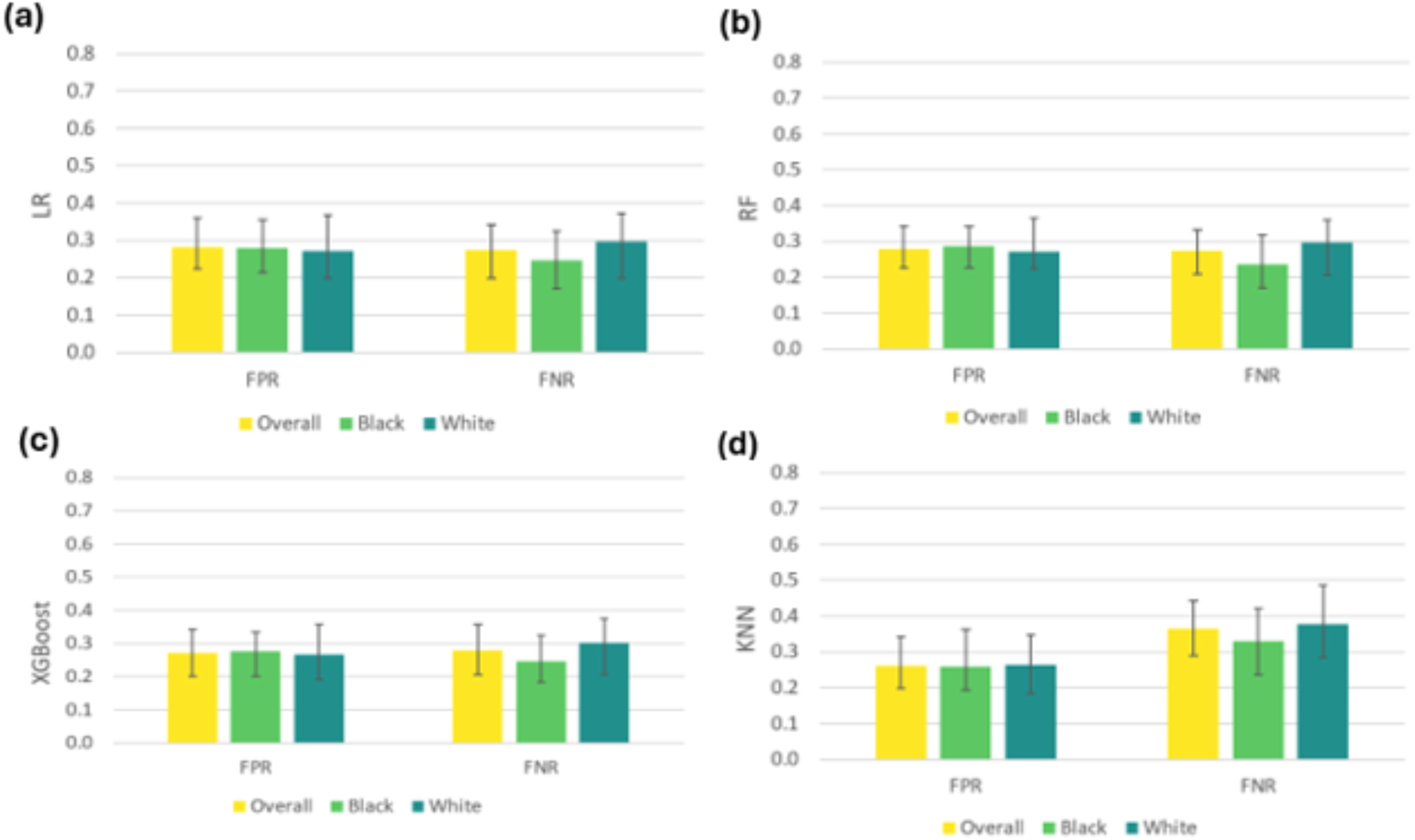
Bias Measures Across Racial Groups. * Bars represent mean performance across 100 random seeds, with error bars indicating 95% confidence intervals. FPR: false positive rates, FNR: false negative rates

### Feature importance

The results of feature importance analysis are shown in Figure 5. Congestive heart failure and inpatient utilization were the two most influential and risk-increasing predictors across models. ED utilization was also demonstrated as a risk-increasing factor. Private insurance emerged as a protective factor. Insured patients showed negative SHAP values across three models.

**Figure 5.**
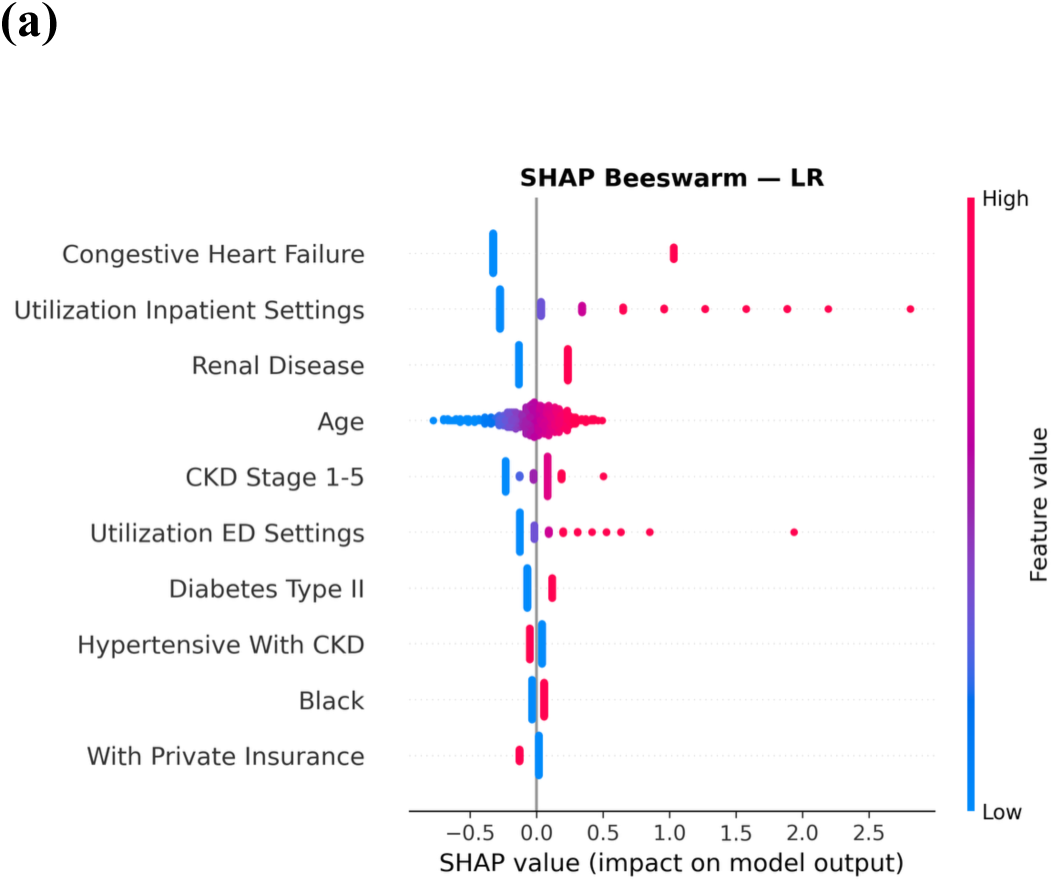
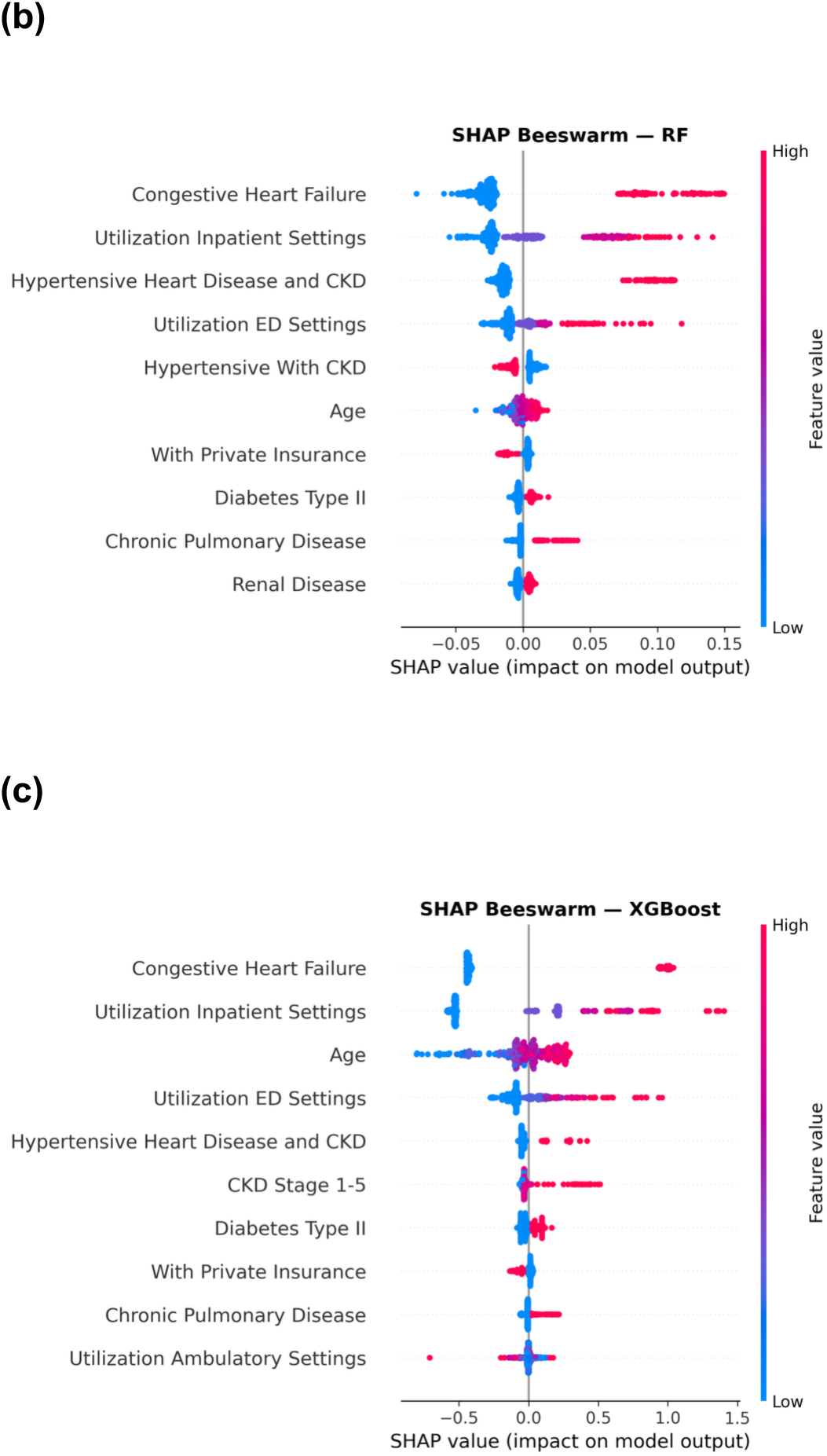
Predictor impact on outcome prediction based on SHAP values displayed using Beeswarm plot (a) LR, (b)RF, and (c) XGBoost.

## Discussion

This study demonstrated the capacity of ML models for predicting 90-day preventable hospital CKD-related admissions using hospital discharge data. Our results showed that LR reached comparable performances with tree-based models (RF, XGBoost), outperforming KNN. Across racial groups, model performances were consistently higher in the Black population than in the white population. Further analysis revealed that all models had a higher FNR in the white population compared to the Black population. These findings can be applied to identify high risk CKD patients and allocate health care resources accordingly.

Models developed in this study were more effective and parsimonious compared with models developed previously to predict preventable CKD-related admissions. Forty five predictors across demographics, comorbidities, hospital utilizations, insurance type, and income data were used to train ML models capable of predicting 90-day preventable admissions. A related previous study, however, used significantly more predictors (e.g. sociodemographic data, clinical characteristics, health care utilization) to predict preventable admissions within a year using the Danish population-level data[11]. With significantly fewer predictors and a shorter window, our models reached comparable performance using the Maryland HCUP data (AuROC: 0.788, AuPRC: 0.416), thus increasing model interpretability and ease of use. The recall at the lowest Youden index threshold was relatively high (above 0.7), resulting in higher F2 scores, which can be used in screening patients with risk of preventable admissions within 90 days.

Applying our models trained using Maryland population data in different racial groups revealed difference in model performance. All models resulted in a higher FNR among white patients then Black patients when predicting preventable CKD admissions (white: 0.297-0.377, Black 0.236-0.33). From the feature importance analysis, we saw that number of utilizations, defined as number of visits in the past year, in both the ED and inpatient settings. Further analysis revealed that white patients had significantly lower utilizations on average than Black patients in the ED (0.75 vs 1.06 visits within one year prior to the ambulatory visit), which can result in a higher FNR. Other potential factors contributing to this finding could include data quality issues (in the ED setting, black patients have better data quality, underutilization in the ambulatory setting)[30].

Our study shows that hospital discharge data can be used to train ML models to predict preventable CKD-related inpatient admissions, which may support risk stratification and healthcare resource planning. These models could help healthcare systems and accountable care organizations identify CKD patients at elevated risk for subsequent preventable admissions and prioritize them for targeted care management. Prior work suggests that CKD-related preventable admissions are not driven by kidney disease alone, but also by comorbidity burden and modifiable clinical factors. Congestive heart failure has been associated with higher odds of preventable ambulatory care-sensitive hospitalization among nursing home residents with CKD[31]. In our study, congestive heart failure was also included as a clinically relevant predictor, supporting its potential role in CKD-related preventable admission risk. However, healthcare systems should be cautious in adopting such models without assessing potential biases, as biased prediction errors may contribute to inequities in health outcomes among CKD patients. The results also have implications for evaluating medical resource use at the state level.

Despite the strength of the findings, limitations should be noted. First, our data was limited to the population of Maryland, thus potentially skewing the training of our models and limiting their generalizability. Additionally, potential data quality biases across racial groups may limit the use of our models in other populations with different racial compositions. Furthermore, as this study relied on HCUP administrative healthcare data, detailed clinical information, such as laboratory values, medication exposures, and gaps in care, was unavailable, limiting our ability to capture the longitudinal complexity of CKD management [32]. Our findings primarily reflect broader associations between patient characteristics and healthcare utilization outcomes, rather than the underlying clinical factors driving these relationships. Finally, the sample size is limited in our dataset, especially for racial groups such as Asians. Hispanic is counted an ethnic group, which can influence the interpretation of our current results.

## Conclusion

This study demonstrated the capacity of ML algorithms in predicting 90-day preventable CKD admission patients using hospital discharge data in outpatient settings while revealing existing biases in model predictions across racial groups (i.e. white, black). Collectively, these findings highlight the clinical relevance of administrative healthcare data for risk stratification and may inform future efforts to improve quality of care and reduce disparities in outcomes among patients with CKD.

## Data Availability

Data produced in the present study are available upon reasonable request to the authors.

## Supplementary Materials

**Supplementary Table 1.**
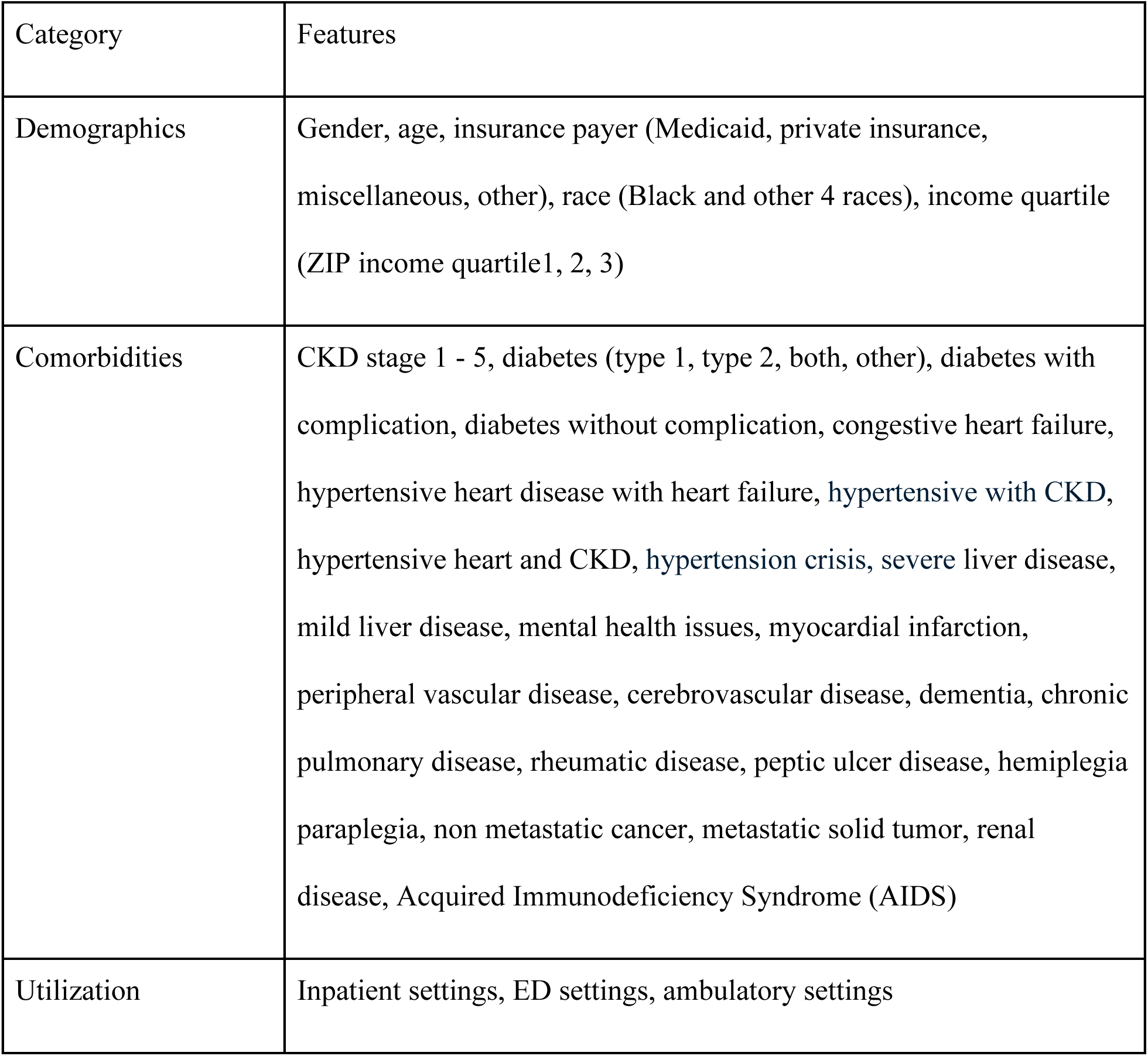
Predictors for 90-day preventable admissions (One-hot encoded variables omitted White, Medicare, and the highest ZIP-code income quartile Q4 as reference categories.)

| Category | Features |
|---|---|
| Demographics | Gender, age, insurance payer (Medicaid, private insurance, miscellaneous, other), race (Black and other 4 races), income quartile (ZIP income quartile1, 2, 3) |
| Comorbidities | CKD stage 1 - 5, diabetes (type 1, type 2, both, other), diabetes with complication, diabetes without complication, congestive heart failure, hypertensive heart disease with heart failure, hypertensive with CKD, hypertensive heart and CKD, hypertension crisis, severe liver disease, mild liver disease, mental health issues, myocardial infarction, peripheral vascular disease, cerebrovascular disease, dementia, chronic pulmonary disease, rheumatic disease, peptic ulcer disease, hemiplegia paraplegia, non metastatic cancer, metastatic solid tumor, renal disease, Acquired Immunodeficiency Syndrome (AIDS) |
| Utilization | Inpatient settings, ED settings, ambulatory settings |

**Supplementary Table 2.** Model performance for predicting 90-day preventable CKD admissions.

|  | **Overall** | **Black** | **White** |
|---|---|---|---|
| **(a) AuROC** |  |  |  |
| **LR** | 0.784(0.768 - 0.799) | 0.797(0.769 - 0.819) | 0.774(0.751 - 0.799) |
| **RF** | 0.787(0.770 - 0.803) | 0.798(0.771 - 0.819) | 0.776(0.752 - 0.804) |
| **XGBoost** | 0.788(0.773 - 0.805) | 0.807(0.749 - 0.857) | 0.776(0.755 - 0.802) |
| **KNN** | 0.753(0.726 - 0.773) | 0.773(0.735 - 0.805) | 0.738(0.707 - 0.767) |
| **(b) AuPRC** |  |  |  |
|  | **Overall** | **Black** | **White** |
| **LR** | 0.397(0.370 - 0.427) | 0.425(0.373 - 0.467) | 0.378(0.331 - 0.418) |
| **RF** | 0.407(0.383 - 0.437) | 0.431(0.379 - 0.476) | 0.391(0.342 - 0.436) |
| **XGBoost** | 0.416(0.377 - 0.459) | 0.467(0.398 - 0.531) | 0.394(0.347 - 0.443) |
| **KNN** | 0.342(0.319 - 0.369) | 0.369(0.329 - 0.406) | 0.329(0.295 - 0.375) |

**Supplementary Table 3.**
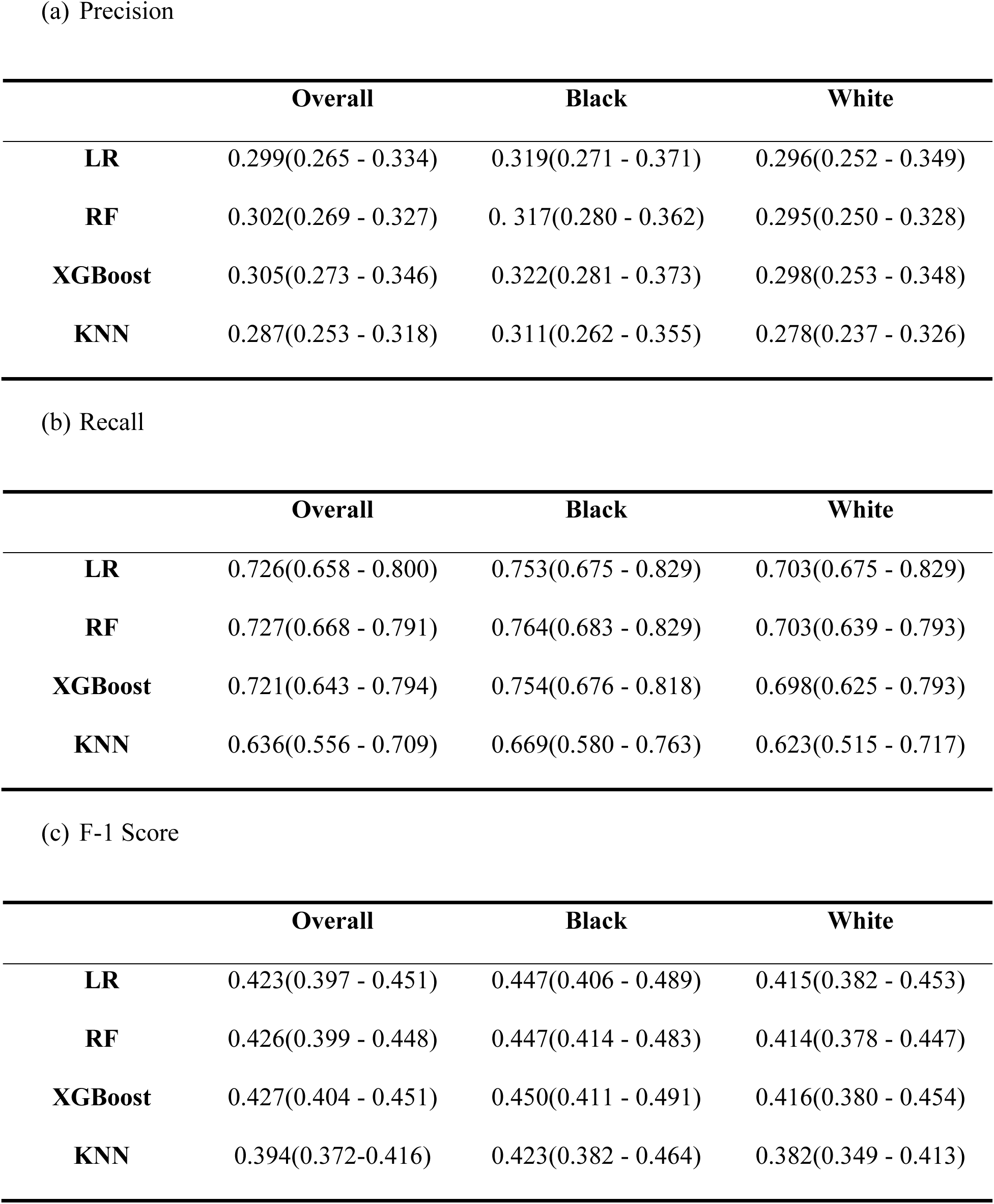
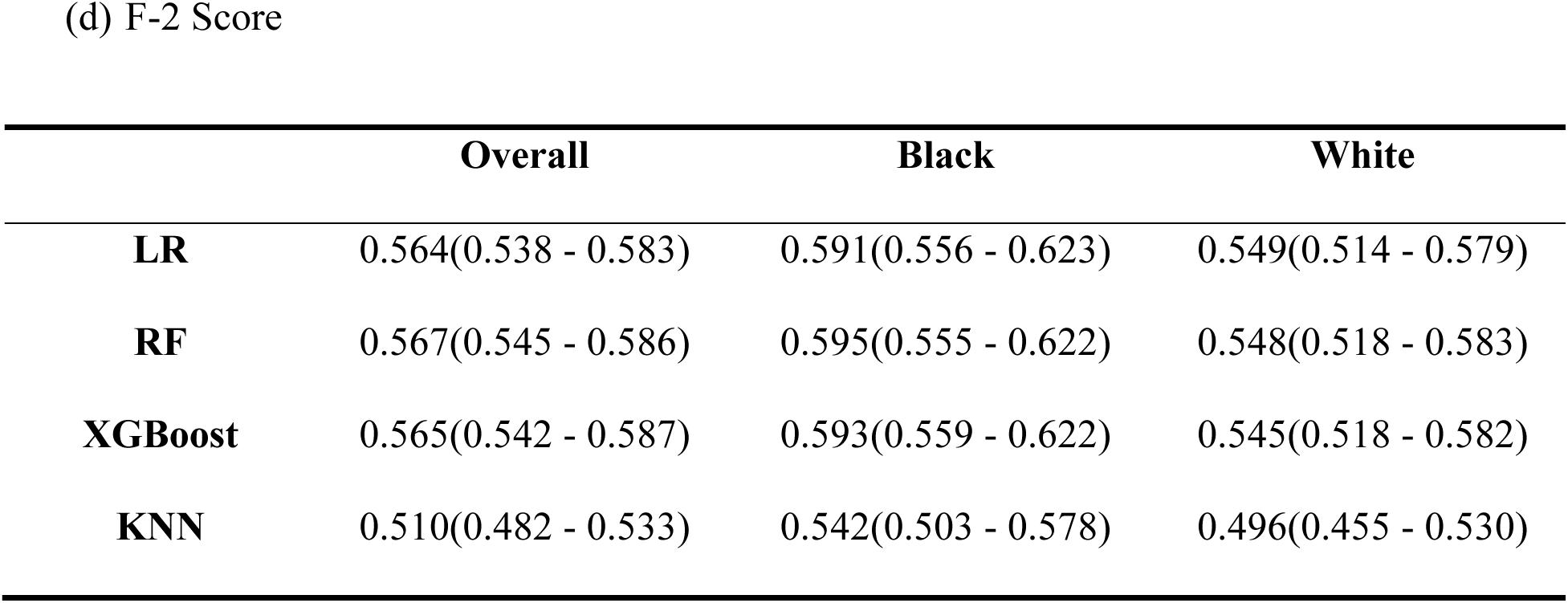
Model Performance Maximizing Youden Index.

|  | **Overall** | **Black** | **White** |
|---|---|---|---|
| (a) Precision |  |  |  |
| **LR** | 0.299(0.265 - 0.334) | 0.319(0.271 - 0.371) | 0.296(0.252 - 0.349) |
| **RF** | 0.302(0.269 - 0.327) | 0. 317(0.280 - 0.362) | 0.295(0.250 - 0.328) |
| **XGBoost** | 0.305(0.273 - 0.346) | 0.322(0.281 - 0.373) | 0.298(0.253 - 0.348) |
| **KNN** | 0.287(0.253 - 0.318) | 0.311(0.262 - 0.355) | 0.278(0.237 - 0.326) |
| (b) Recall |  |  |  |
|  | **Overall** | **Black** | **White** |
| **LR** | 0.726(0.658 - 0.800) | 0.753(0.675 - 0.829) | 0.703(0.675 - 0.829) |
| **RF** | 0.727(0.668 - 0.791) | 0.764(0.683 - 0.829) | 0.703(0.639 - 0.793) |
| **XGBoost** | 0.721(0.643 - 0.794) | 0.754(0.676 - 0.818) | 0.698(0.625 - 0.793) |
| **KNN** | 0.636(0.556 - 0.709) | 0.669(0.580 - 0.763) | 0.623(0.515 - 0.717) |
| (c) F-1 Score |  |  |  |
|  | **Overall** | **Black** | **White** |
| **LR** | 0.423(0.397 - 0.451) | 0.447(0.406 - 0.489) | 0.415(0.382 - 0.453) |
| **RF** | 0.426(0.399 - 0.448) | 0.447(0.414 - 0.483) | 0.414(0.378 - 0.447) |
| **XGBoost** | 0.427(0.404 - 0.451) | 0.450(0.411 - 0.491) | 0.416(0.380 - 0.454) |
| **KNN** | 0.394(0.372-0.416) | 0.423(0.382 - 0.464) | 0.382(0.349 - 0.413) |
| (d) F-2 Score |  |  |  |
|  | **Overall** | **Black** | **White** |
| **LR** | 0.564(0.538 - 0.583) | 0.591(0.556 - 0.623) | 0.549(0.514 - 0.579) |
| **RF** | 0.567(0.545 - 0.586) | 0.595(0.555 - 0.622) | 0.548(0.518 - 0.583) |
| **XGBoost** | 0.565(0.542 - 0.587) | 0.593(0.559 - 0.622) | 0.545(0.518 - 0.582) |
| **KNN** | 0.510(0.482 - 0.533) | 0.542(0.503 - 0.578) | 0.496(0.455 - 0.530) |

**Supplementary Table 4.** Bias Measures for Predicting Preventable Admission.

|  | **Overall** | **Black** | **White** |
|---|---|---|---|
| (a) LR |  |  |  |
| **FPR** | 0.281(0.223 - 0.361) | 0.279(0.213 - 0.354) | 0.271(0.197 - 0.368) |
| **FNR** | 0.274(0.199 - 0.342) | 0.247(0.171 - 0.325) | 0.297(0.199 - 0.372) |
| (b) RF |  |  |  |
|  | **Overall** | **Black** | **White** |
| **FPR** | 0.278(0.227 - 0.343) | 0.286(0.225 - 0.342) | 0.271(0.224 - 0.366) |
| **FNR** | 0.273(0.209 - 0.332) | 0.236(0.171 - 0.317) | 0.297(0.207 - 0.361) |
| (c) XGBoost |  |  |  |
|  | **Overall** | **Black** | **White** |
| **FPR** | 0.272(0.201 - 0.343) | 0.277(0.202 - 0.335) | 0.266(0.191 - 0.358) |
| **FNR** | 0.279(0.206 - 0.357) | 0.247(0.182 – 0.324) | 0.302(0.207 - 0.375) |
| (d) KNN |  |  |  |
|  | **Overall** | **Black** | **White** |
| **FPR** | 0.262(0.199 - 0.341) | 0.259(0.193 - 0.363) | 0.263(0.182 - 0.347) |
| **FNR** | 0.364(0.290 - 0.444) | 0.330(0.237 - 0.420) | 0.377(0.283 - 0.485) |

**Supplementary Table 5.** Selected hyperparameters for four models.

|  | Tuning method | Selected hyperparameters | Selection AuROC |
|---|---|---|---|
| **Logistic Regression** | Grid Search | {'C': 0.1, 'penalty': 'l1'} | 0.793 |
| **Logistic Regression** | Bayesian Optimization | {'C': 0.0981228189480366, 'penalty': 'l1'} | 0.792 |
| **Random Forest** | Grid Search | {'max_depth': 10, 'min_samples_split': 10, 'n_estimators': 500} | 0.797 |
| **Random Forest** | Bayesian Optimization | {'n_estimators': 334, 'max_depth': 13, 'min_samples_split': 4, 'min_samples_leaf': 8} | 0.797 |
| **XGBoost** | Grid Search | {'learning_rate': 0.01, 'max_depth': 3, 'n_estimators': 500, 'subsample': 0.6} | 0.8 |
| **XGBoost** | Bayesian Optimization | {'max_depth': 4, 'learning_rate': 0.014978173620818836, 'n_estimators': 233, 'subsample': 0.7424334028017774, 'colsample_bytree': 0.9092737981534443} | 0.8 |
| **KNN** | Grid Search | {'metric': 'manhattan', 'n_neighbors': 50, 'weights': 'distance'} | 0.752 |
| **KNN** | Bayesian Optimization | {'n_neighbors': 44, 'weights': 'distance', 'metric': 'manhattan'} | 0.752 |

**Supplementary Table 6.**
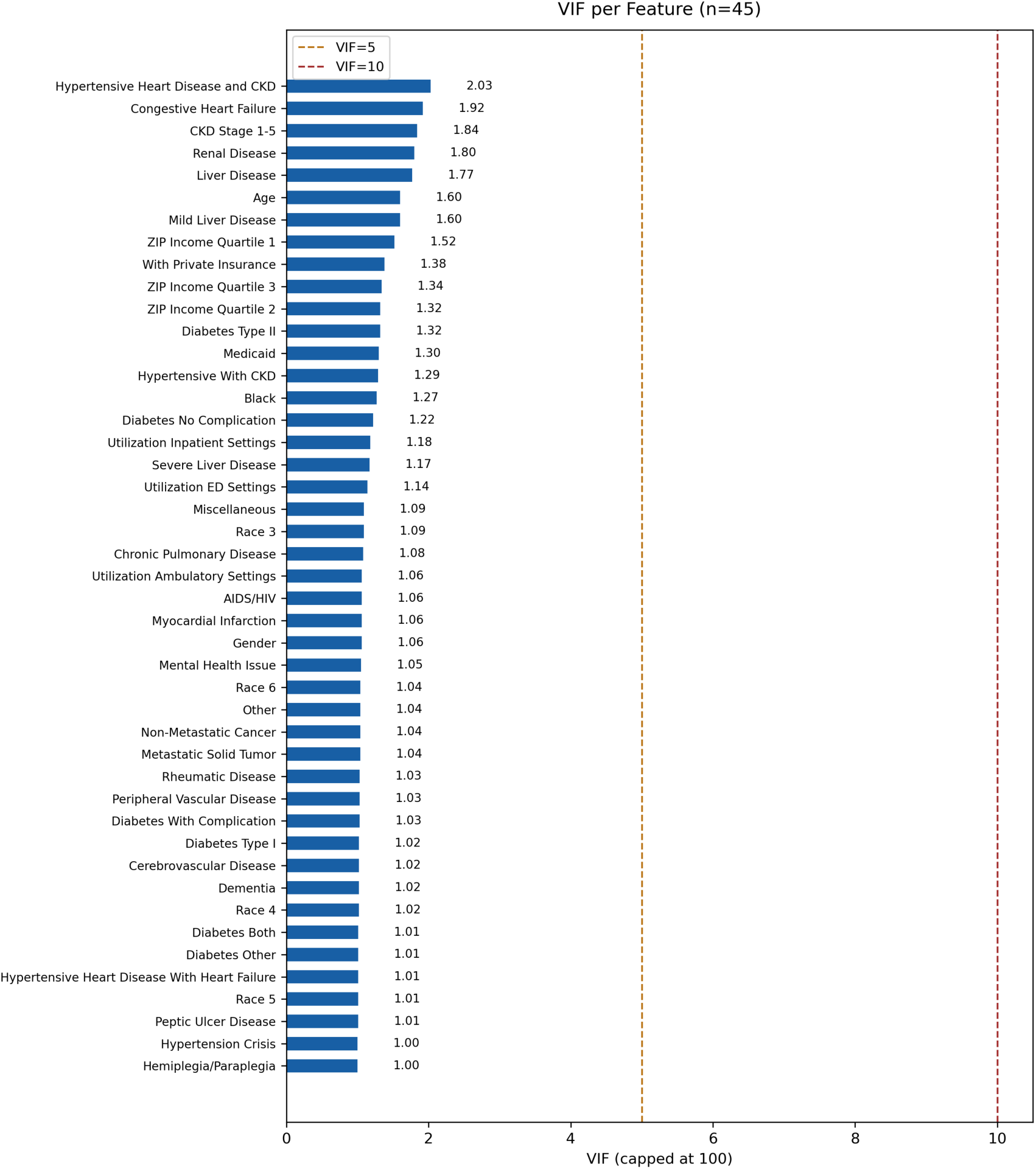
Variation Inflation Report among features.

| Feature | VIF (capped at 100) |
|---|---|
| Hypertensive Heart Disease and CKD | 2.03 |
| Congestive Heart Failure | 1.92 |
| CKD Stage 1-5 | 1.84 |
| Renal Disease | 1.80 |
| Liver Disease | 1.77 |
| Age | 1.60 |
| Mild Liver Disease | 1.60 |
| ZIP Income Quartile 1 | 1.52 |
| With Private Insurance | 1.38 |
| ZIP Income Quartile 3 | 1.34 |
| ZIP Income Quartile 2 | 1.32 |
| Diabetes Type II | 1.32 |
| Medicaid | 1.30 |
| Hypertensive With CKD | 1.29 |
| Black | 1.27 |
| Diabetes No Complication | 1.22 |
| Utilization Inpatient Settings | 1.18 |
| Severe Liver Disease | 1.17 |
| Utilization ED Settings | 1.14 |
| Miscellaneous | 1.09 |
| Race 3 | 1.09 |
| Chronic Pulmonary Disease | 1.08 |
| Utilization Ambulatory Settings | 1.06 |
| AIDS/HIV | 1.06 |
| Myocardial Infarction | 1.06 |
| Gender | 1.06 |
| Mental Health Issue | 1.05 |
| Race 6 | 1.04 |
| Other | 1.04 |
| Non-Metastatic Cancer | 1.04 |
| Metastatic Solid Tumor | 1.04 |
| Rheumatic Disease | 1.03 |
| Peripheral Vascular Disease | 1.03 |
| Diabetes With Complication | 1.03 |
| Diabetes Type I | 1.02 |
| Cerebrovascular Disease | 1.02 |
| Dementia | 1.02 |
| Race 4 | 1.02 |
| Diabetes Both | 1.01 |
| Diabetes Other | 1.01 |
| Hypertensive Heart Disease With Heart Failure | 1.01 |
| Race 5 | 1.01 |
| Peptic Ulcer Disease | 1.01 |
| Hypertension Crisis | 1.00 |
| Hemiplegia/Paraplegia | 1.00 |

**Supplementary Figure 1.**
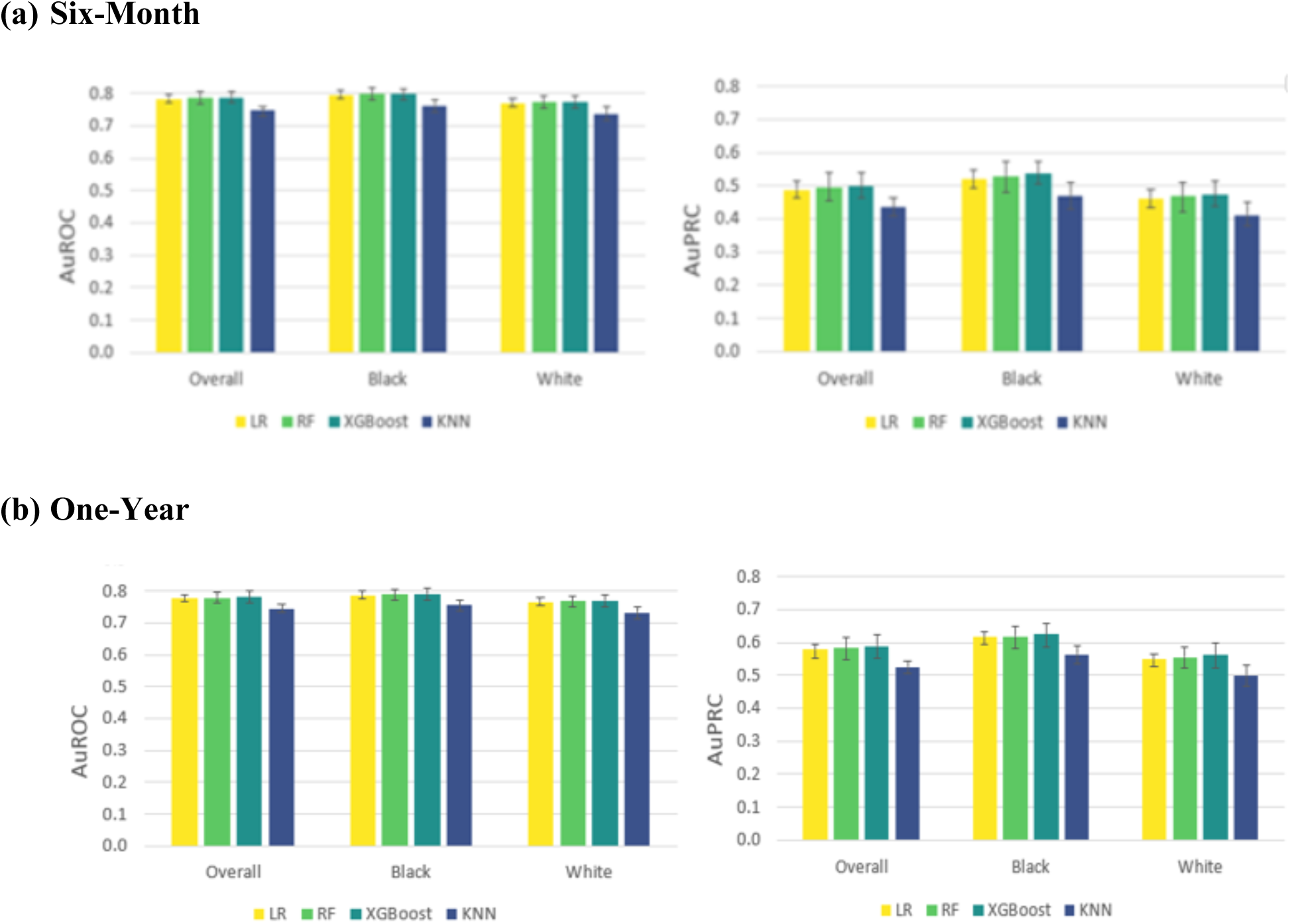
Predicting Preventable Admissions Using Different Timeframes.

**Supplementary Figure 2.**
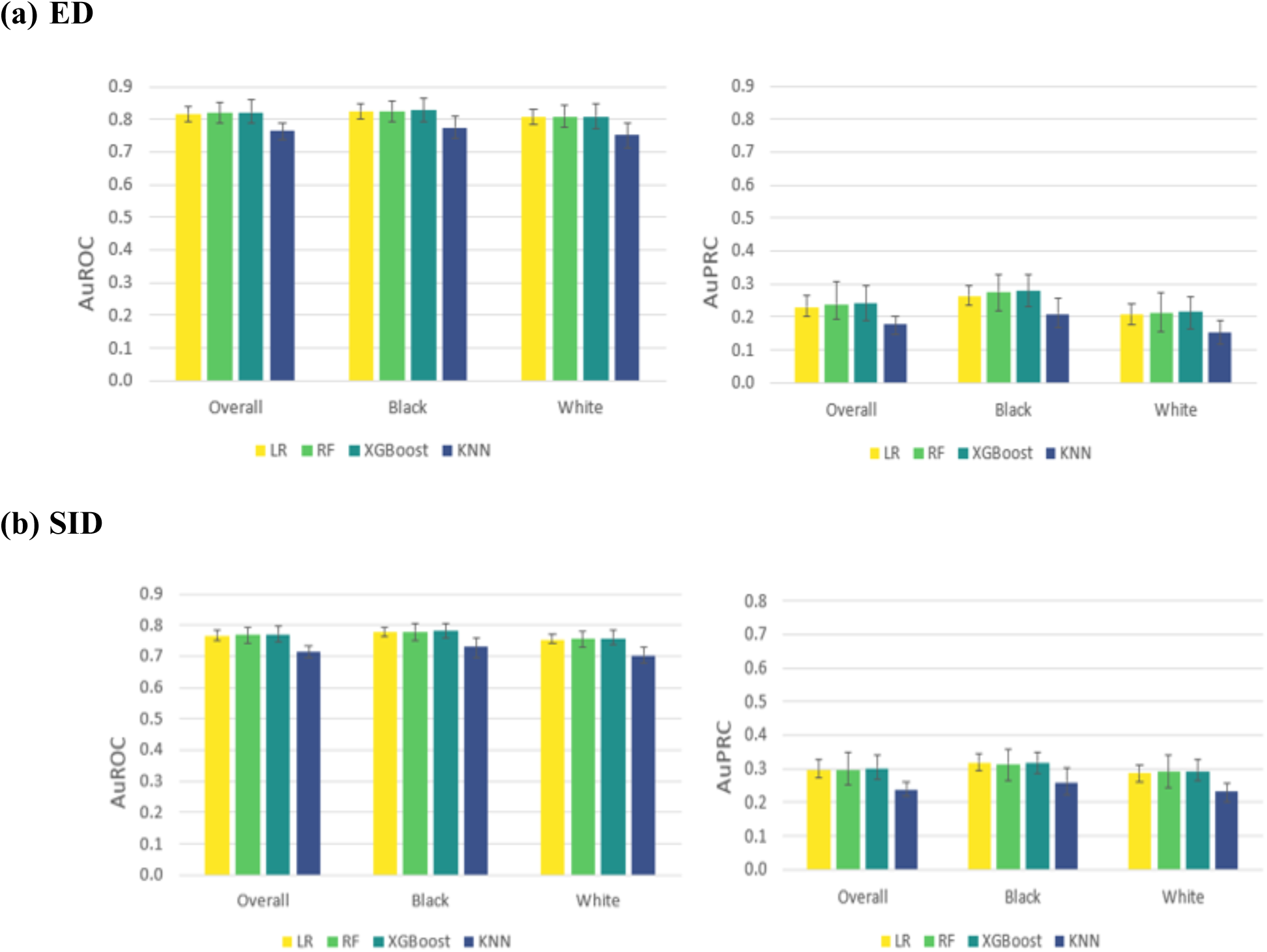
Predicting 90-Day Preventable ED Admissions and Hospitalization.

## Conflict of interest

The authors have no conflict of interest to disclose with this study.

## Author contribution

Conceptualization: YCL, CP, HK, DB

Data curation: YCL, HK

Methods Development: YCH, YH, YCL

Data Analysis: YCH, YH, YCL

Data Visualization: YCH, YH, YCL

Manuscript Writing: YCH, YH

Manuscript Editing and Review: YCL, CP, HK, DB

## Notes

### Competing Interest Statement

The authors have declared no competing interest.

### Author Declarations

HCUP data are deidentified and exempt from Institutional Review Board (IRB) reviews.

